# Transformer based deep learning denoising of single and multi-delay 3D Arterial Spin Labeling

**DOI:** 10.1101/2023.04.24.23288718

**Authors:** Qinyang Shou, Chenyang Zhao, Xingfeng Shao, Kay Jann, Karl G. Helmer, Hanzhang Lu, Danny JJ Wang

## Abstract

**Purpose:** To present a Swin Transformer-based deep learning (DL) model for denoising of single-delay and multi-delay 3D arterial spin labeling (ASL) and compare its performance with convolutional neural network (CNN) methods.

**Methods:** Swin Transformer and CNN-based spatial denoising models were developed for single-delay ASL. The models were trained on 59 subjects (104 scans) and tested on 44 subjects (57 scans) from 3 different vendors. Spatiotemporal denoising models were developed using another dataset (6 subjects, 10 scans) of multi-delay ASL. A range of input conditions was tested for denoising single and multi-delay ASL respectively. The performance was evaluated using similarity metrics, spatial signal-to-noise ratio (SNR) and quantification accuracy of cerebral blood flow (CBF) and arterial transit time (ATT).

**Results:** Swin Transformer outperformed CNN-based networks, whereas pseudo-3D models showed better performance than 2D models for denoising single-delay ASL. The similarity metrics and image quality (SNR) improved with more slices in pseudo-3D models, and further improved when using M0 as input but introduced greater biases for CBF quantification. Pseudo-3D models with 3 slices as input achieved optimal balance between SNR and accuracy, which can be generalized to different vendors. For multi-delay, spatiotemporal denoising models had better performance than spatial-only models with reduced biases in fitted CBF and ATT maps.

**Conclusions:** Swin Transformer DL models provided better performance than CNN methods for denoising both single and multi-delay 3D ASL data. The proposed model offers flexibility to improve image quality and/or reduce scan time for 3D ASL to facilitate its clinical use.

## 1. Introduction

Arterial Spin Labeling (ASL) is an appealing MRI technique for studying brain function since it is entirely non-invasive and is able to quantify hemodynamic parameters such as cerebral blood flow (CBF) and arterial transit time (ATT). However, a major limitation of ASL is its relatively low sensitivity and signal-to-noise ratio (SNR) due to the small fraction (1-2%) of the labeled arterial blood and its T1 relaxation during the measurement. The past decade has seen a range of technical advances including pseudo-Continuous ASL (pCASL), background suppression (BS) and 3D acquisitions in conjunction with high magnetic field strength that have facilitated the clinical translation of ASL[1]. However, it remains challenging to reliably apply ASL to the diagnosis and management of individual patients, especially in aging populations. Denoising algorithms based on spatial and/or temporal filtering have been proposed to improve the robustness and SNR of ASL, often at the cost of reduced imaging coverage and sharpness[2], [3].

Deep learning (DL) techniques have recently shown great promises and versatility in various fields including medical imaging[4]. As the most widely used DL technique, convolutional neural networks (CNN), such as U-net[5] and ResNet[6] is able to automatically capture the hierarchical and complex features of the input image and can identify, classify, and quantify patterns in medical images[7], [8]. During recent years, CNN-based denoising algorithms have been proposed for ASL to suppress noise/artifacts[7], [9], which are potentially superior to standard denoising algorithms in terms of preserving the original image resolution and sharpness. Kim et al. [9] first introduced a CNN-based network for ASL denoising with local and global pathways. Xie et al. [7] developed a Dilated Wide Activation Network (DWAN) for denoising ASL images which has been applied to 2D pulsed ASL (PASL) data in persons with mild cognitive impairment and Alzheimer’s disease[7], [10]. Gong et al.[8] introduced multi-contrast input including ASL and M0 images to the network to further improve the performance for denoising 3D pCASL data.

Transformer[11] is a newly developed DL model that adopts the mechanism of self-attention, differentially weighting the significance of each part of the input data to provide long-range dependencies especially for time series. Transformer has been successfully applied in the fields of natural language processing [11], computer vision[12], and recently expanded to medical image processing[13]. In particular, Vision Transformer[12] was first proposed to directly apply Transformer on images, which can integrate both short and long range features with the multi-head self-attention mechanism. Swin Transformer[14] was proposed to serve as a general-purpose backbone for computer vision, which calculates attention in local windows to improve efficiency for different image scales. SwinIR[15] was built upon Swin Transformer and combined with Convolution layers to perform lower-level image restoration task like super-resolution or denoising. These features of SwinIR were desirable for medical imaging as medical images always have different contrasts in different image regions, which may be useful in image enhancement tasks. In addition, Transformer-based network preserves local features compared to CNN, which may have shortcoming of spatial blurring[15], [16].

The purpose of the present study is to present a Swin Transformer-based DL model for denoising single-delay and multi-delay 3D ASL data and compare its performance with CNN-based methods. We compared three model backbones, a Swin Transformer-based network (SwinIR) and two CNN-based networks (DWAN and ResNet), with different 2D and pseudo-3D input conditions. We evaluated the performance by measuring similarity with the reference, SNR in the perfusion images, and systematic bias in quantification of CBF. Finally, spatiotemporal denoising models were developed for denoising multi-delay ASL data.

## 2. Methods

### 2.1 Problem formulation

ASL is commonly acquired with multiple repetitions including pairs of control and label images to obtain an average perfusion image with sufficient SNR and a proton density image (M0) for quantification. Here we propose a flexible scheme that is based on performing DL denoising on each perfusion image calculated with a single pair of control and label images, which can then be averaged to obtain a perfusion image with higher SNR. Alternatively, this scheme allows fewer repetitions with shorter scan time to achieve a perfusion image with sufficient SNR. In this study, we focused on single and multi-delay 3D pCASL data acquired on 3T MRI scanners of different vendors. 3D acquisition is the recommended form of ASL acquisition by the consensus paper[1] and offers higher SNR compared to 2D acquisitions.

A summary of the 6 datasets used in this study is summarized in Table 1. The participants (total N=116, age=60±18, 74 females) were generally healthy without major neurologic/psychiatric disorder or severe systemic disease, and provided written informed consents. The participants of dataset 1, 3-5 were part of the MarkVCID consortium study [17]at the University of Southern California (USC), Massachusetts General Hospital and Johns Hopkins University, while the participants of dataset 2 and 6 underwent MRI scanning at USC.

**Table 1.** Details of the datasets used in this study, including the number of subjects and cases, patient age and gender, MRI scanner vendors, ASL parameters including labeling duration (LD) and post labeling delay (PLD), image resolution, matrix size, number of repetitions (L/C pairs) and the number of testing cases.

|  | Dataset 1 | Dataset 2 | Dataset 3 | Dataset 4 | Dataset 5 | Dataset 6<br>(Multi-delay) |
| --- | --- | --- | --- | --- | --- | --- |
| Vendor | Siemens<br>Prisma | Siemens<br>Prisma | Siemens<br>Prisma | Philips<br>Achieva | GE Discovery<br>750W | Siemens<br>Prisma |
| N subjects | 55(15 males) | 25(11 males) | 10 (4 males) | 10 (3 males) | 10 (4 males) | 6 (5 males) |
| Age | 69±7 | 38±20 | 65±10 | 69±8 | 71±9 | 35±13 |
| N scans | 98 | 48 | 10 | 10 | 10 | 10 |
| LD/PLD (ms) | 1500/2000 | 1800/2000 | 2000/2500 | 2000/2500 | 2000/2500 | 1800/500-2500 |
| Resolution<br>(mm<sup>3</sup>) | 2.5×2.5×2.5 | 2.5×2.5×2.5 | 3.4×3.4×4 | 3.4×3.4×4 | 3.4×3.4×4 | 2.5×2.5×2.5 |
| Matrix size | 96×96×48 | 96×96×40 | 64×64×36 | 80×80×36 | 128×128×36 | 96×96×40 |
| Repetitions<br>(L/C pairs) | 7 | 9 | 8 | 5 | 1 | 9/9/9/9/3 (for<br>delay 1-5) |
| N test case | 9 | 18 | 10 | 10 | 10 | 3-fold cross<br>validation |
| *Some subjects have 2 scans |  |  |  |  |  |  |

For single-delay ASL, the training data were chosen from the first 2 datasets acquired on 3T Siemens Prisma scanner using background suppressed 3D GRASE (gradient and spin echo) pCASL. The trained DL models were first tested on unseen data from the same cohorts (datasets 1-2) as the training data, and were also independently tested on 3D pCASL data acquired on Siemens Prisma, Philips Achieva and GE Discovery 750 3T scanners respectively (datasets 3-5) with different imaging parameters[18]. Each scan in the dataset was organized as a 4-D matrix of perfusion images with first three dimensions as image volume and the last dimension as the repetition. For the training stage, each individual repetition was served as the input and the average of all repetitions was used as the reference.

In multi-delay ASL, several perfusion images with different PLDs were acquired and used to fit parametric maps such as CBF and ATT[19]. For multi-delay ASL model training and evaluation, we used a separate multi-delay dataset (dataset 6) with 3D GRASE pCASL acquisition and five PLDs of 500/1000/1500/2000/2500ms on a Siemens 3T Prisma scanner. We performed denoising for the perfusion image of a single repetition for each PLD respectively, which were then used to fit for CBF and ATT maps. In this study, we first validated the method on single-delay ASL data for spatial denoising, and then extended the proposed method to multi-delay ASL for spatiotemporal denoising.

### 2.2 Network architecture

In this work, a Swin Transformer-based network SwinIR[15] and two state-of-the-art CNN network architectures, DWAN[7] and ResNet[20] were investigated. The SwinIR network architecture for denoising is shown in Figure 1. Input images were first processed by a convolution block for shallow feature extraction, which was followed by 6 residual Swin Transformer blocks for deep feature extraction. Each Swin Transformer block was composed of 6 consecutive Swin Transformer layers with a convolutional block at the end. Compared to vision Transformer, Swin Transformer uses a shifted window approach, where the input image is divided into several non-overlapping windows and the self-attention is calculated within each window. Window shifting is made in consecutive Swin Transformer layers to provide connections between these windows. The output of the Swin Transformer blocks is finally processed by a convolution layer to perform the image denoising task. For ResNet, the input image was processed by 20 residual convolution blocks without batch normalization according to Lim et al.[20]. As a benchmark for CNN-based network for ASL denoising, DWAN network was implemented using the same structure reported in the original paper by Xie et al.[7].

**Figure 1.**
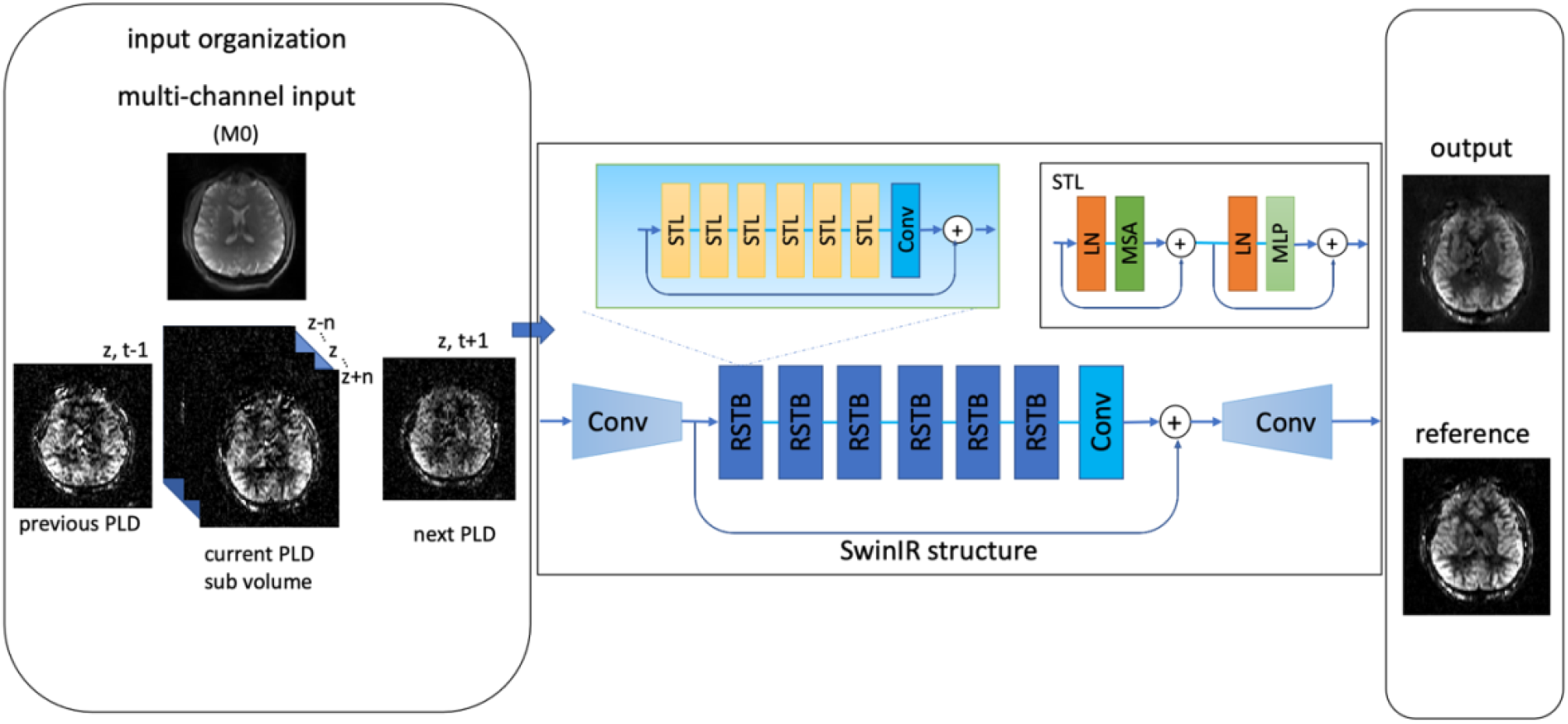
The framework for the ASL denoising task. The input to the model was an image slice combined with several other channels, which are images from spatially adjacent slices, or image from temporally adjacent PLDs. The SwinIR structure was composed of a shallow feature extraction module (convolution layer), a deep feature extraction module (residual Swin Transformer blocks (RSTB)) and an image restoration module (convolution layer). (STL: Swin Transformer layer, LN: layer normalization, MSA: multi-head self-attention, MLP: multi-layer perceptron)

For each of the backbones, the baseline model uses a 2D structure which takes a single slice as the input and outputs a single slice. Given that the perfusion image is a 3D volume, adjacent slices may provide useful information for denoising. Pseudo-3D method is a way to incorporate adjacent slices in the input[21]. Instead of a single slice, a sub-volume of N slices (odd number) which contains the center slice, and N-1 adjacent slices are used as the input, which feeds more spatial information to the model. In this study, several pseudo-3D methods were investigated (N=1, 3, 5, 7, N=1 means standard 2D model). We also investigated multi-modality input conditions, where the M0 image was included as an additional channel to the input[8]. Since the M0 image is usually acquired with ASL for quantification, this method does not require a separate scan or coregistration. A detailed setting of the experiments is summarized in Supplementary Table S1. The 2D with M0 condition took one slice of perfusion image and the M0 image of the same slice. The M0-with-3-slice condition took 3 slices from the perfusion images and the center slice of the corresponding M0 image as input.

### 2.3 Training and testing procedures

All networks were implemented with Pytorch 1.12 [22] and python 3.7 and trained on a lambda cluster with NVIDIA 3090 GPU. The ASL control and label images were motion corrected using SPM12 and pairwise subtracted to get the perfusion images. The perfusion images were averaged to get the reference for each scan. Since there might be large artifact or severe head motion occurred across the perfusion series, a quality control step was performed to ensure the training data did not include outliers. Perfusion images with mean signal greater or less than mean±2 standard deviations of the perfusion image time series were considered outliers and excluded from the training data. (On average, less than 1 time frame of each scan was excluded from the training data). The top and bottom 10% of the slices were cropped since they didn’t have enough SNR and/or often contained artifacts. The data were divided into 8:1:1 for training, validation and testing respectively. Each perfusion image and its corresponding reference were considered an input-reference pair, resulting in a total of 104 training scans with 28443 slices and 15 validation scans with 4018 slices. A sub-volume training strategy was used where a 48×48×N sub-volume randomly chosen from the image instead of the whole was used as the input. Data augmentation strategies included random image rotation within the range of -60 to 60 degrees and range flip along x direction. A combined loss with ℓ_1_ and structural similarity index (SSIM) loss, where

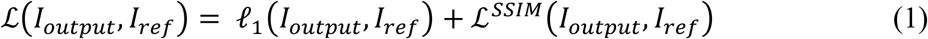

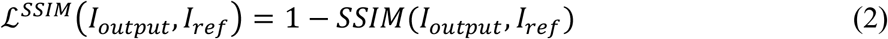

was used as the loss function to preserve both local features and perceptual image quality. ADAM optimizer was used with initial learning rate of 10^−3^. The learning rate was reduced if the performance had not improved after 20 epochs. Batch size was 16 for all model settings. Each model was trained for 500 epochs, and the parameters from the model that achieved the best performance on the validation dataset was recorded.

### 2.4 Model evaluation

We tested the model performance on the test data as described in Table1. The perfusion images were first normalized to the range of 0 and 1 and fed into the model, then the output was scaled back to the input scale according to the pre-recorded minimum and maximum values of the original image after processing. The predicted CBF maps were calculated using the predicted perfusion image and the original M0 image with the following equation:

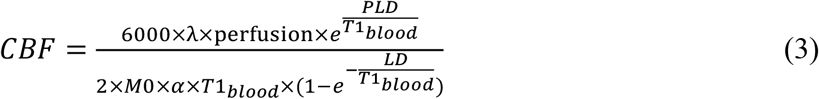

where T1_blood_ was 1650ms, λ is the brain blood partition coefficient and assumed to be 0.9[1], *α* is labeling efficiency was set to 0.735 based on simulation accounting for 2 background suppression (BS) pulses (Siemens and Philips data), LD and PLD were the labeling duration and post labeling delay and were set to the actual scan parameters. For GE ASL data, α was 0.6 with 4 BS pulses and the M0 image was extrapolated from a saturation recovery image with a brain T1 of 1200ms and a 2000ms delay following the saturation pulse[23].

The performance of the models was evaluated in three aspects. First, the similarity metrics including peak signal-to-noise ratio (PSNR) and structural similarity index (SSIM) were measured between the predicted CBF maps and the reference. These metrics were calculated for each repetition and then averaged for the scan. Paired t tests were used to compare performance metrics between each pair of 3 models for each input condition respectively.

Second, spatial SNR was calculated in the gray matter mask by the definition in Feinberg et al. [24], with the mean of the signal averaged across all repetitions, divided by the standard deviation of the difference image between even-number-averaged and odd-number-averaged perfusion images. The spatial SNR was calculated for perfusion images of either part of or all repetitions (2, 4 and all repetitions) respectively. This SNR calculation provided an image quality measurement without a required reference. Third, processed perfusion images of either part of or all repetitions (2, 4 and all repetitions) were averaged to produce an averaged CBF map. Whole brain, gray matter and white matter CBF values were calculated within masks for processed method and reference to evaluate the systematic bias introduced by the models. Mean difference between reference and predicted CBF values along with its 95% confidence interval was calculated. Gray matter (GM), white matter (WM), and whole brain masks were segmented from M0 image using SPM12. Since existing literature reported test-retest variability of ASL scans on the order of 10% [25]–[29], we consider CBF and ATT biases less than 10% within the normal variation range and clinically acceptable. The intraclass correlation coefficient (ICC) of absolute agreement between the prediction and the reference was calculated to test the accuracy of the predicted values.

For independent evaluation in 3D pCASL data of 3 vendors, the models trained on datasets 1 and 2 were directly applied to the test dataset 3-5 without fine-tuning. Note the similarity metrics of SSIM and PSNR cannot be calculated as the GE scanner provided only one perfusion image which was the average of 3 repetitions. The SNR was calculated by the mean of signal in the gray matter divided by the standard deviation in the white matter for GE perfusion images.

### 2.5 Model extension to multi-delay ASL

The aforementioned DL denoising schemes on single-delay ASL data were extended to multi-delay ASL data with 3-slice and temporally adjacent perfusion images (pseudo-4D, see Figure 1) as input for spatiotemporal denoising. For multi-delay ASL, it is important to not only improve the SNR for each perfusion image, but also to preserve the temporal relationship across different PLDs. Therefore, we included one temporal dimension as the input to constrain the dynamic relationship between PLDs to improve the model performance in estimating quantitative maps. In this experiment, we acquired a separate multi-delay 3D GRASE pCASL dataset (dataset 6 in Table 1) with 5 delays (500, 1000, 1500, 2000, 2500ms) and 9/9/9/9/3 repetitions for each delay respectively, fewer repetitions for last PLD were acquired due to time limits. Similar to the previous experiments, each individual perfusion image was used as input and the average of all timepoints of that delay was used as the reference. The three model backbones, DWAN, ResNet and SwinIR were tested with either spatial input (center + 2 adjacent slices) or spatiotemporal input (center + 2 adjacent slices and 2 adjacent PLDs). We padded in the temporal dimension to get enough input for the first and last PLDs. We used a 3-fold cross validation for the training and in each fold 1/3 of the subjects would be left for the test group so that every subject can be used once for evaluation. For evaluations, Predicted CBF and ATT maps were fitted from DL processed 5-delay perfusion images with repetitions of 1,1,1,2,2, which would result in a 5-minute scan.

Reference CBF and ATT maps were fitted using the 5-delay perfusion images averaged with all repetitions. The similarity metrics between the output and reference quantitative maps were calculated, and differences in mean CBF and ATT value in whole brain, GM and WM masks were calculated. The similarity metrics (SSIM, PSNR), and quantification accuracy using mean difference in masks and ICC were similarly calculated as described in single-delay experiment.

## 3. Results

### 3.1 Single delay ASL results on dataset 1 and 2

The average inference time was 0.22, 0.23 and 0.33 seconds per slice for DWAN, ResNet and SwinIR.

Figure 2 shows the DL denoised perfusion images with different model backbones and input settings on a representative subject. The input image is an individual perfusion image calculated by subtraction of one control-label pair and the subtitles indicate the model from which the input was processed. Compared to the input image, DL denoised images had higher SNR and better gray and white matter contrast. Figure 3 shows an example of the prediction by a 3-slice SwinIR model with 12 slices covering the cortex. The reference was averaged by all repetitions of original perfusion images, and the prediction was averaged by 2, 4 and all repetitions of denoised perfusion images respectively. The SNR of output perfusion images was the highest after DL processing and averaging all repetitions which improved the SNR by approximately 2-fold compared to the reference image.

**Figure 2.**
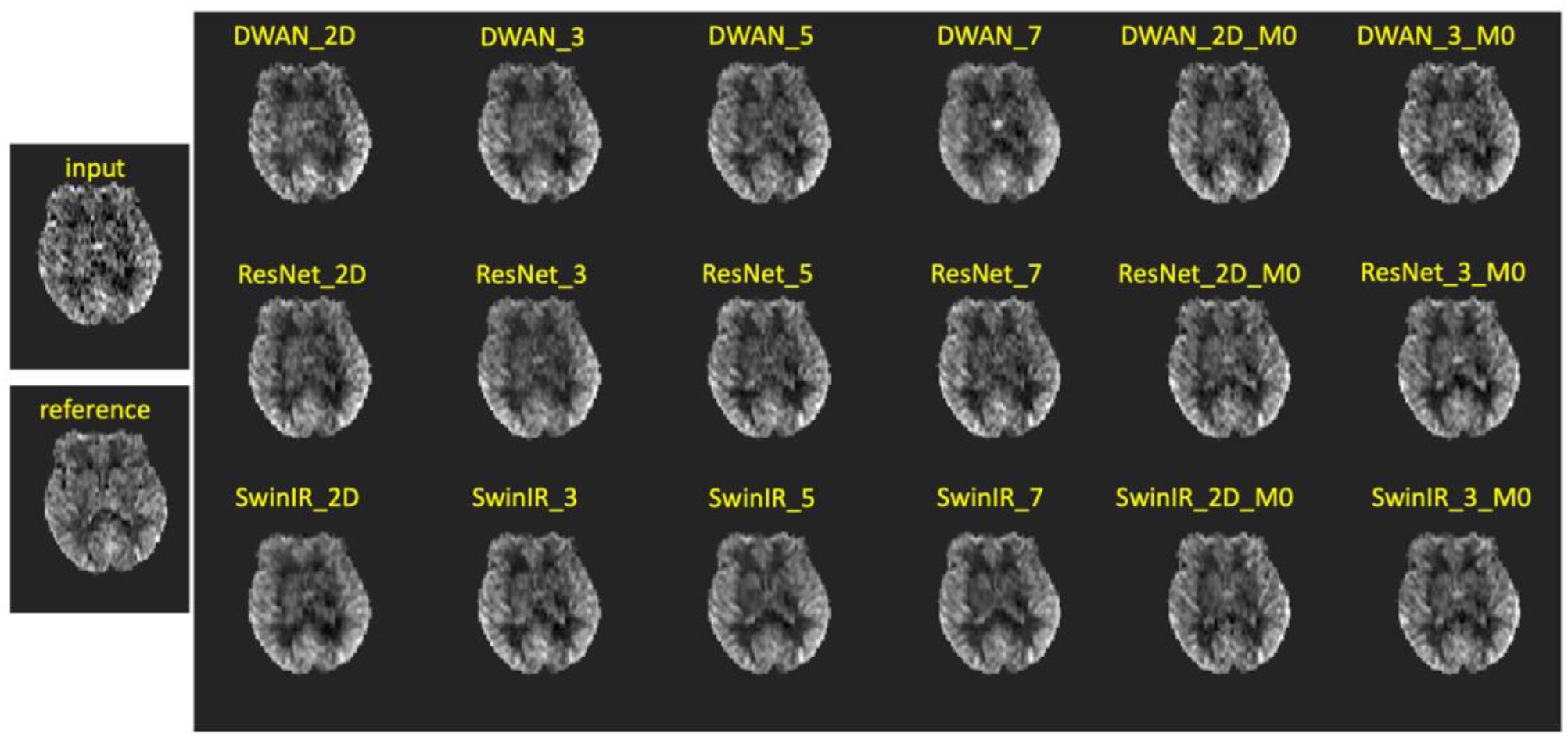
Input, reference, and prediction perfusion images processed by the deep learning models of different settings. The DL processed image has higher SNR compared to the input image and high similarity to the reference.

**Figure 3.**
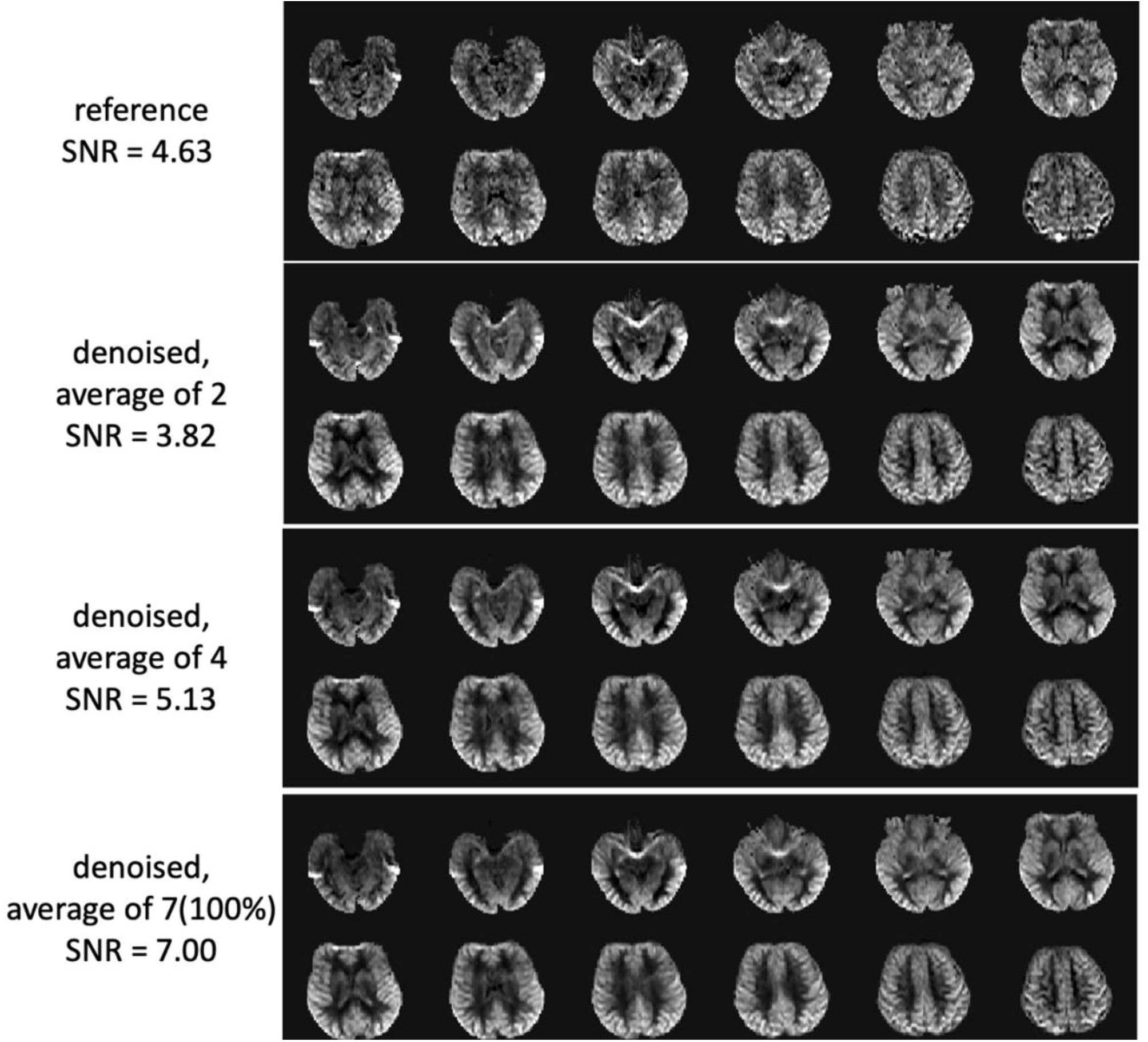
Example with larger coverage of a representative subject. The reference is averaged by all input perfusion images. Each input perfusion image was denoised by the DL model and averaged by different portions of the time points (2, 4 and all). More averaging will result in higher SNR.

Figure 4 shows the comparison of similarity metrics for different model backbones and settings respectively. Within the same backbone model the SSIM and PSNR improved as the slice channel increased, where the improvement from 5 slice to 7 slice was marginal. The models with the M0 image as additional input outperformed all model settings with only perfusion images as the input. Among the three model backbones, SwinIR outperformed ResNet and DWAN in all model settings (p<0.001), except for the 2D model, the SSIM of ResNet was slightly higher than that of SwinIR (p<0.01). Supplementary Figure S1 shows the SNR of the output perfusion image with different averages. The trend for different model settings was similar to that of the similarity metrics, where increased input channels increased SNR and including the M0 image as additional channel improved SNR the most. With more averages of DL denoised perfusion images, the SNR was further improved. The reference image with the average of all time points achieved SNR of 4.840±1.627 (dashed red line), while a comparable and higher SNR can be achieved with only ∼50% the data points. If all the data points were used in averaging, the SNR can be improved by about 2-fold. The details of all model results were included in supplementary Table S2 and Table S3.

**Figure 4.**
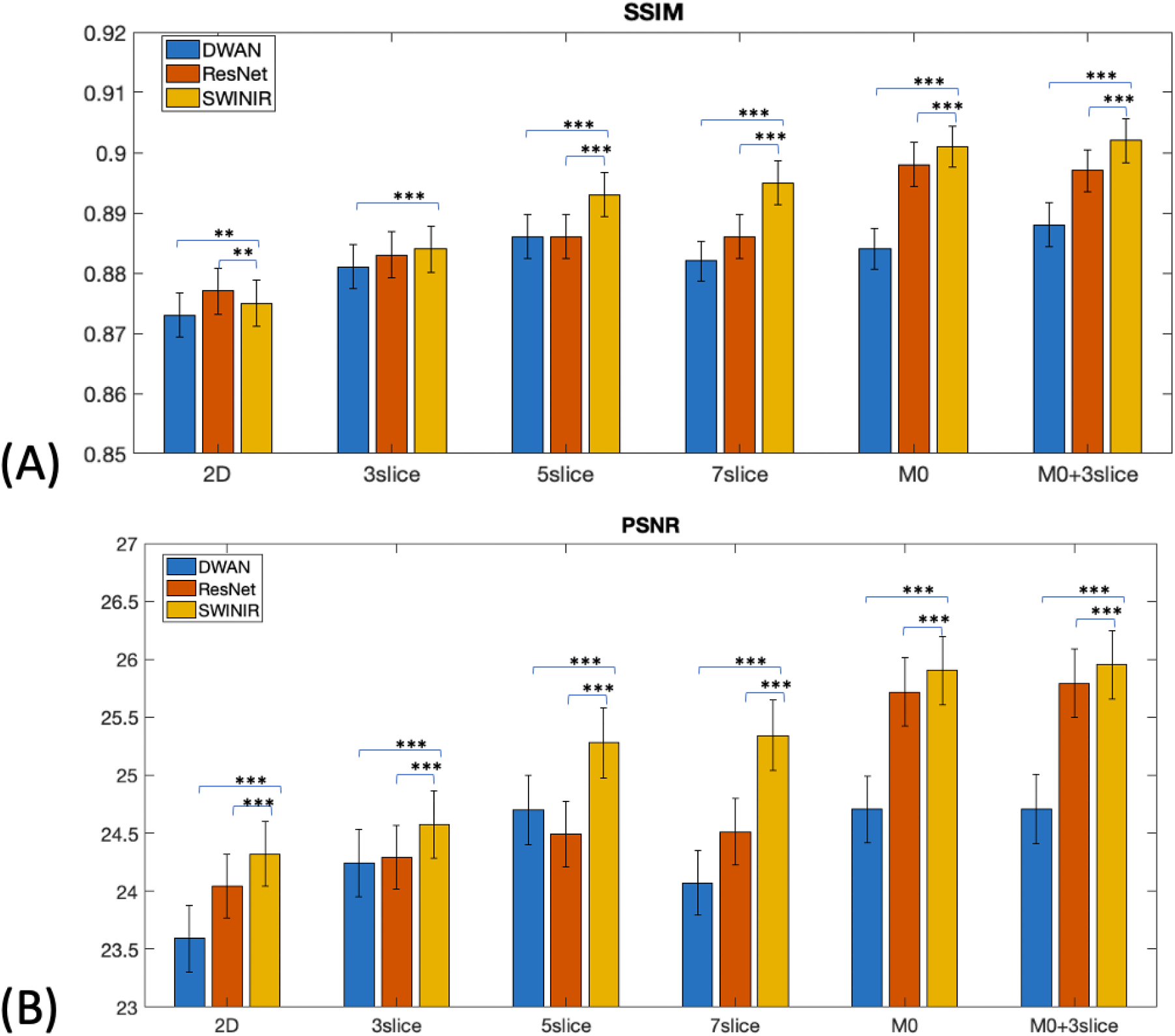
Comparison of the SSIM, PSNR of different model backbones and input settings. For both SSIM and PSNR, with more adjacent slices to the input channels, the performance improves. Adding a M0 channel will result in the largest improvement. The significance of the difference was indicated on the bar plot (*: p<0.05, **: p<0.01, ***: p<0.001)

Supplementary Figure S2 shows the scatter plot for all 3 baseline models. It can be observed that with more averages, the predicted CBF values became closer to the reference. Figure 5(A) shows the mean difference of CBF values for whole brain, GM, and WM using all repetitions for different model settings respectively. In Figure 5(B), it can be seen that for models that didn’t include M0 as input, the mean CBF differences were relatively small (within 10%), while the models that included M0 as input had significantly higher biases in WM CBF compared to other input settings for all 3 model backbones. For SwinIR models without M0 as input, the mean value in the GM was slightly increased by less than 5%, while the mean value in WM was decreased by less than 10% except for 7-slice pseudo-3D input. The detailed results of the biases and the ICCs for the prediction and the reference are shown in supplementary Table S4. Based on the above results, we chose pseudo-3D input with 3 slices as the optimal input condition for independent evaluation on 3D pCASL data of 3 different vendors below, based on the balance between increased SNR and minimal bias for CBF quantification as well as computational complexity.

**Figure 5.**
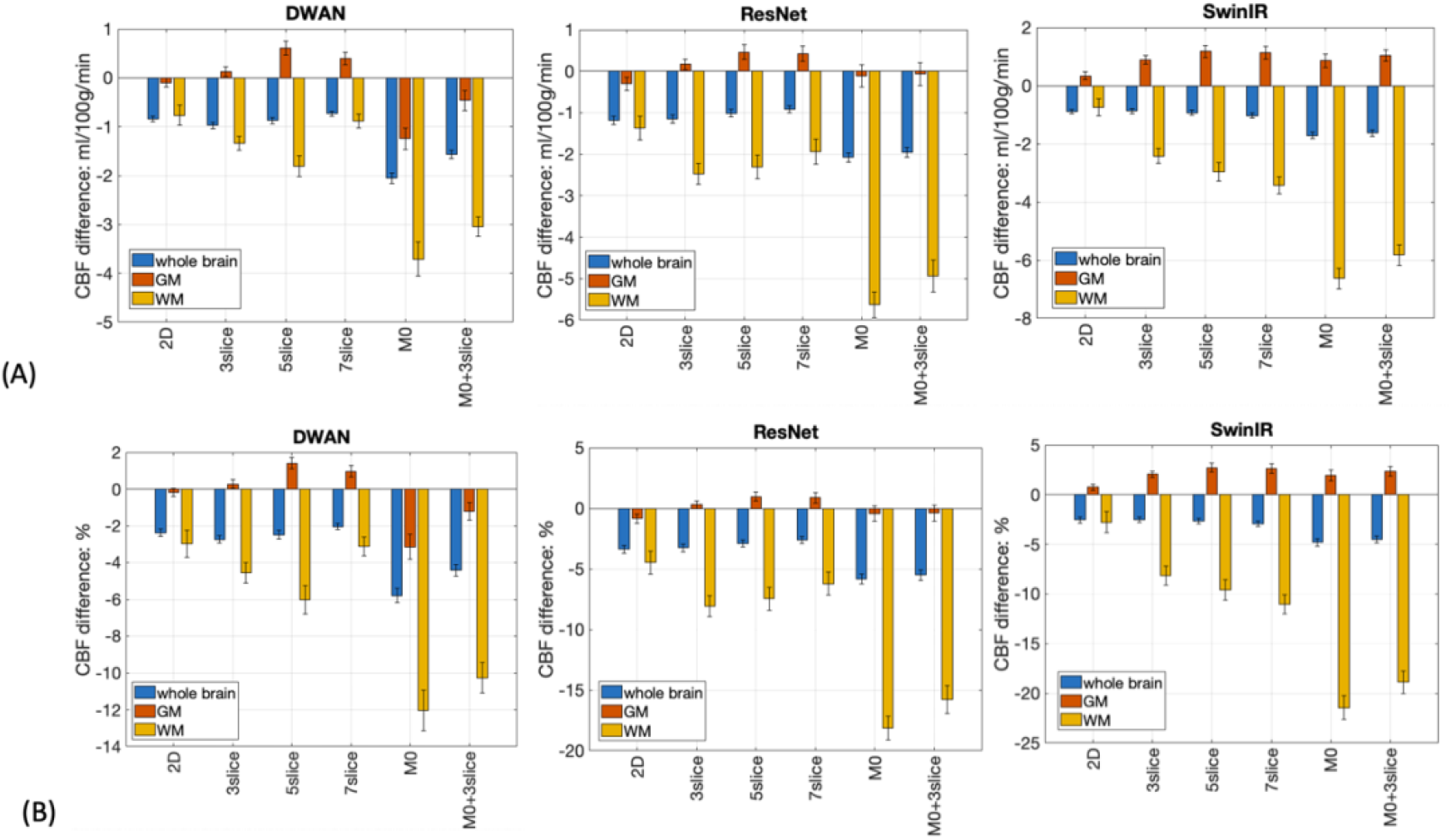
Mean difference of CBF values in whole brain, gray matter, and white matter (relative values (A) and in percentage (B)) for different model backbones and input settings.

### 3.2 Single delay ASL results on dataset 3-5

Figure 6 shows the denoising performance for the perfusion images from different cohorts acquired on 3 MR vendors. For all three vendors, there was an increase in SNR for the perfusion images. For GE data, since the original data was already the average of 3 repetitions with high SNR (individual repetition data were not provided by the vendor), the improvement by DL denoising was marginal. Table 2 shows the details of improvement in SNR of different model backbones with 3 slice input for 3 vendors respectively. All three models were able to improve SNR over the input. ResNet achieved the best performance for Philips data, SwinIR achieved best performance for the Siemens data, and DWAN performed best on GE data.

**Figure 6.**
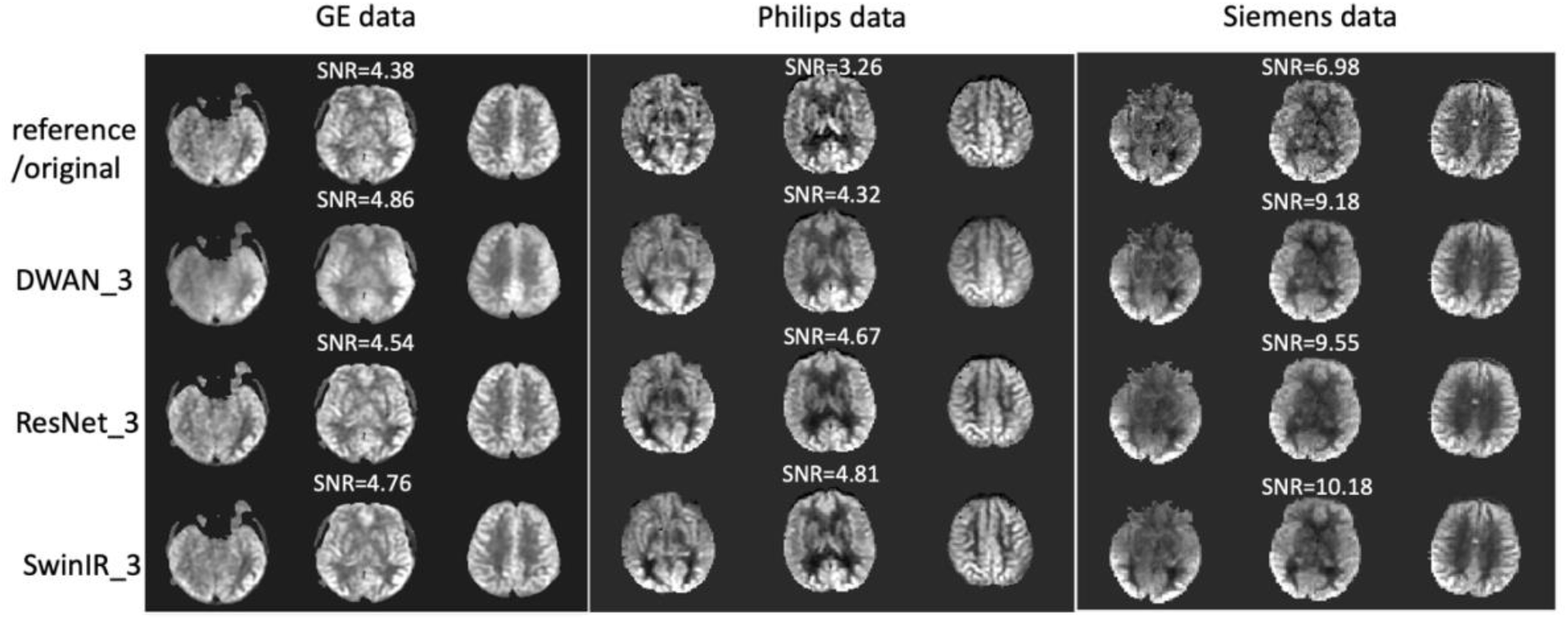
Denoising performance of 3 models for 3 representative cases from independent testing cases from different vendors. For GE data, the original image is the input to the networks. For the other 2 vendors, the reference images are averaged by all input repetitions. SNR of the images are shown above each case.

**Table 2.** SNR performance for different vendors with different proportion of averages. The best performance across different models was shown in bold.

| Model | Philips |  |  | Siemens |  |  | GE |
| --- | --- | --- | --- | --- | --- | --- | --- |
|  | SNR 2 | SNR 4 | SNR all | SNR 2 | SNR 4 | SNR all | SNR all |
| ResNet_3 | **3.509±1.077** | **5.005±1.607** | **5.582±1.701** | **3.109±1.308** | 4.278±1.935 | 6.379±2.705 | 4.398±0.457 |
| SwinIR_3 | 3.108±1.229 | 4.476±1.594 | 4.988±1.580 | 3.104±1.281 | **4.391±2.017** | **6.485±2.789** | 4.561±0.210 |
| DWAN_3 | 3.001±1.069 | 4.244±1.554 | 4.714±1.576 | 2.716±1.141 | 3.843±1.777 | 5.754±2.632 | **4.725±0.589** |
| Input | 2.171±0.861 | 3.101±1.107 | 3.524±1.143 | 2.088±0.884 | 2.948±1.352 | 4.359±1.975 | 4.105±0.504 |

Supplementary Table S5 shows the bias analysis for three different vendors. For Siemens data, SwinIR achieved the highest ICC in whole brain and GM CBF values, while ResNet achieved the highest ICC in the WM. For Philips and GE data, all three backbones achieved similar performance except that DWAN had a large bias in WM CBF (>10%).

### 3.3 Multi-delay ASL results

Figure 7 shows denoising results of a representative multi-delay dataset. The SNR of the perfusion image of each PLD was improved with all models. Figure 8 shows the fitted CBF and ATT maps from two representative subjects. The input CBF and ATT maps were fitted from the 5-delay perfusion images with fewer repetitions (1, 1, 1, 2, 2) and the reference maps were fitted from the 5-delay perfusion images with all repetitions (9, 9, 9, 9, 3). The DL models denoised the input perfusion image of each delay and the CBF and ATT maps were fitted from the denoised 5-delay perfusion images. It can be seen that after DL denoising, the fitted CBF maps and ATT maps achieved higher SNR compared to those calculated from the input images. The models with an extra temporal denoising dimension showed better performance in the WM compared to the spatial only denoising models. The red arrows in Figure 8 indicate spurious high ATT values in input images which were suppressed in spatiotemporal denoised images using 3 PLD input.

**Figure 7.**
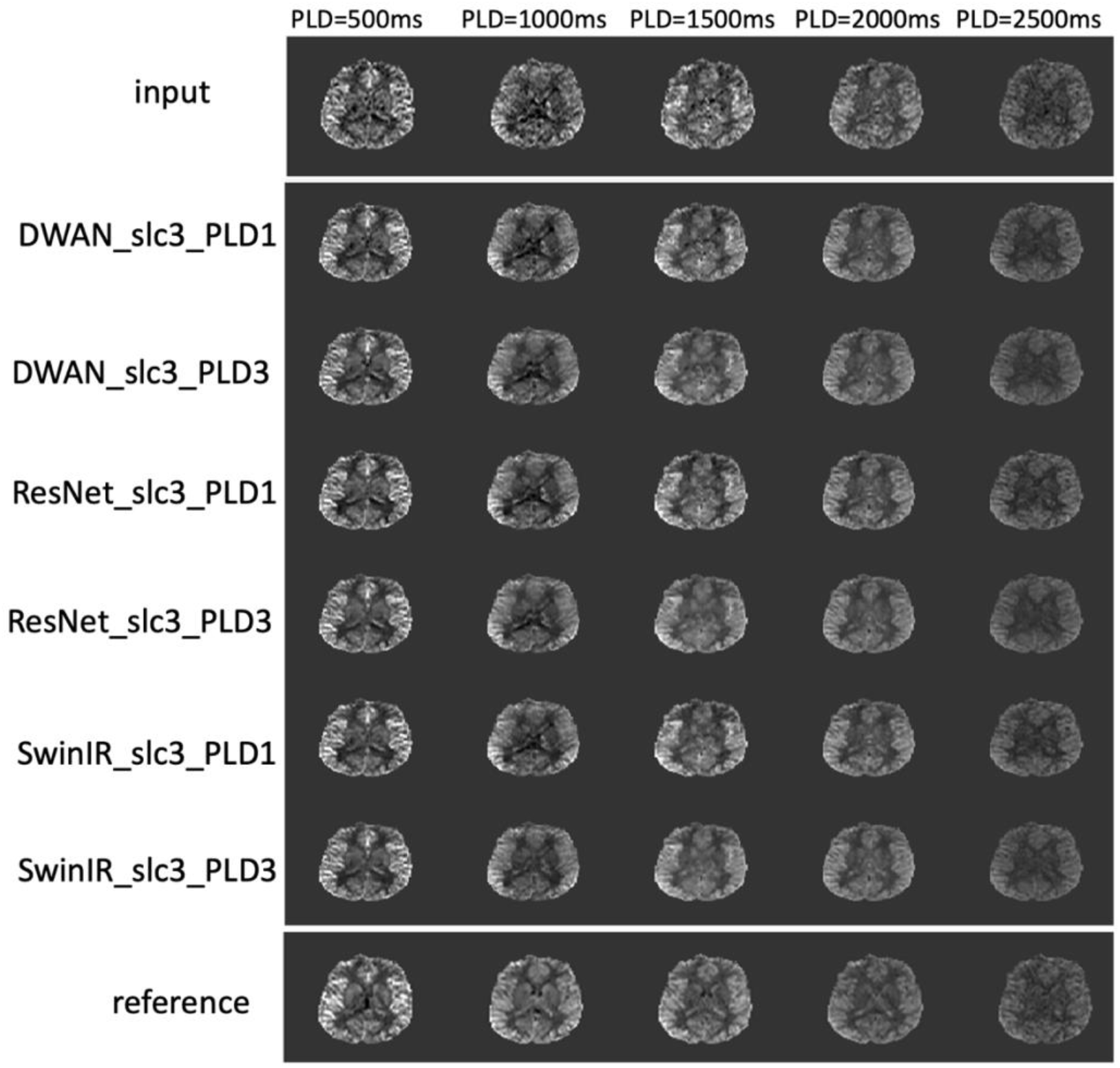
An example of the multi-delay dataset. Input perfusion images were averaged with 1,1,1,2,2 repetitions for PLD of 500, 1000, 1500, 2000 and 2500ms. Reference perfusion images were averaged with 9,9,9,9,3 repetitions for PLD of 500, 1000, 1500, 2000 and 2500ms. The denoised perfusion images were DL predictions with the input, which show improves SNR for each PLD.

**Figure 8.**
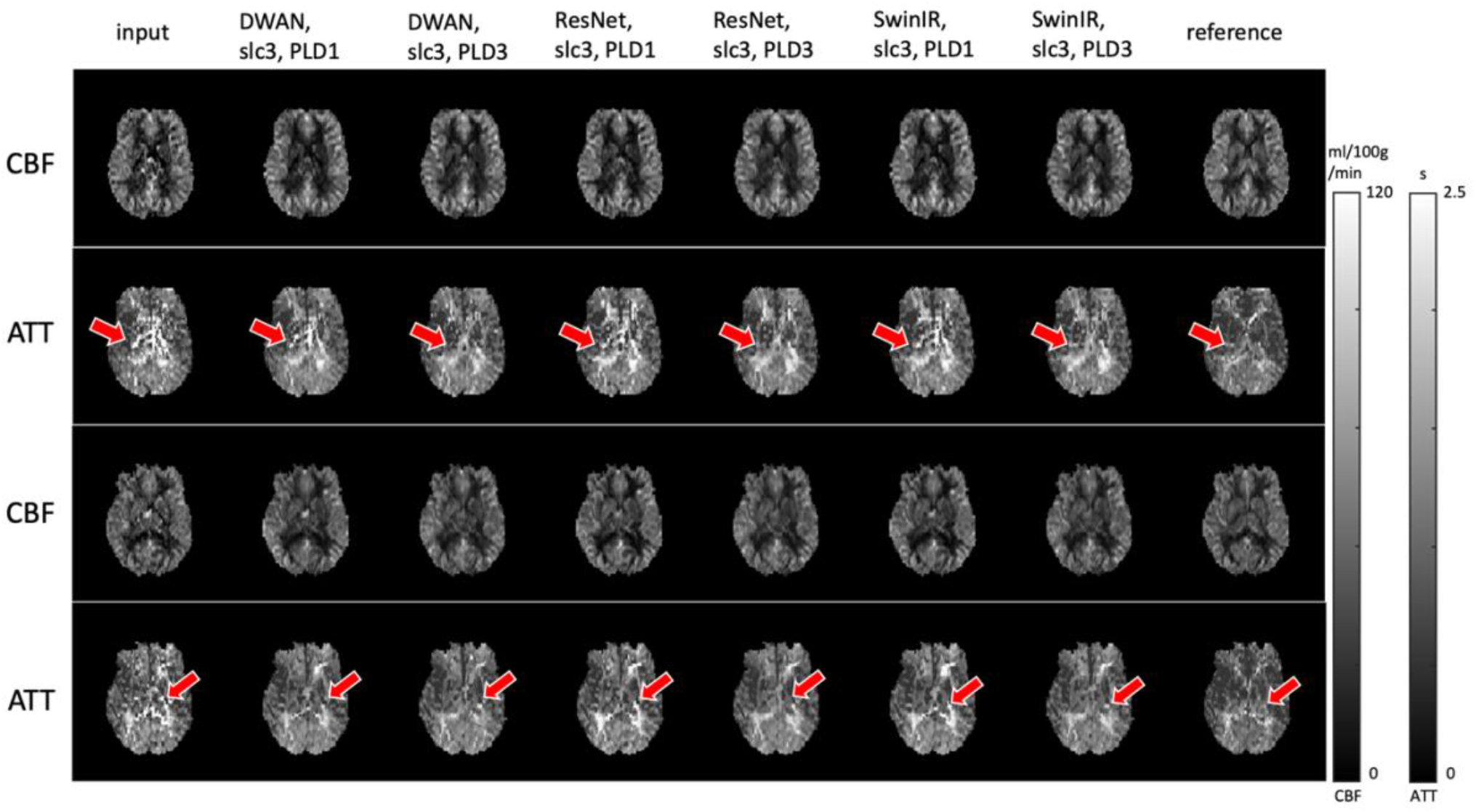
Fitted CBF and ATT maps for two representative subjects. Red arrows show a spot where an error in fitting occurs due to spike in the input, which was resolved in spatiotemporal denoising models, but not resolved in spatial only models.

Supplementary Figure S3 shows the similarity and bias analysis for the CBF and ATT maps of the denoising models. The results were averaged for all test subjects across the 3-fold cross validation. Supplementary Figure S3 (A) shows that the spatial denoising models and spatiotemporal denoising models achieved similar performance for the CBF maps, nevertheless the spatiotemporal models achieved significantly higher similarity for the fitted ATT maps. In all experimental settings, SwinIR outperformed ResNet and DWAN in the quantitative metrics (SSIM and PSNR) of both CBF and ATT maps. Details of similarity metrics can be found in supplementary Table S6. Supplementary Figure S3 (B) shows the differences in the CBF and ATT values between the denoised image and reference. For the CBF values, all models achieved relatively small bias (the largest difference in GM and WM CBF was 5.97% and 6.29% respectively). For the ATT values, all models showed small biases in gray matter (within 10%), but the spatiotemporal models showed better performance with small ATT biases in the WM (∼ 2%), whereas the spatial only models showed a difference of ∼8% in WM ATT. Details of bias analysis can be found in supplementary Table S7.

## 4. Discussion

### 4.1 Study innovation

In this study, we presented a Swin Transformer-based DL method to perform ASL denoising and compared its performance with CNN-based methods. There are 4 innovations in our method: 1) To the best of our knowledge, this is the first study applying Transformer for denoising ASL images and one of first few studies for denoising medical images[30]–[32]; 2) We employed a flexible strategy to denoise each single perfusion image which can be averaged according to the specific need for either reducing total scan time and/or achieving image enhancement; 3) We trained DL models using 3D pCASL data acquired on Siemens 3T scanner and tested the models on 3D pCASL data acquired on different cohorts and MRI vendor platforms, as well as on a multi-delay 3D pCASL dataset. All these ASL acquisitions are state-of-the-art and consistent with consensus recommendation; 4) Last but not the least, three state-of-the-art network backbones, ResNet, DWAN and SwinIR were adapted for ASL denoising with a range of input conditions including 2D and 3 pseudo-3D input conditions (N=3,5,7) and multi-model input with both perfusion and M0 images. Furthermore, we extended the model with pseudo-4D input to perform spatiotemporal denoising on multi-delay ASL data.

### 4.2 Optimal input conditions and model backbone

In this study, several input conditions were investigated including pseudo-3D and combination with M0 image. Compared to the proposed pseudo-3D strategy, direct 3D models have the following limitations. First, 3D convolution layers have more parameters compared to their 2D counterparts, which will result in a larger model size. Second, training a 3D network will need more working memory than 2D models since the 3D input size is larger, and this will reduce feasible batch size given the limitation of GPU memory. Third, 3D model training and inferencing will take longer than 2D models. For these reasons, we proposed a 2D network with proper pseudo-3D input which provides more flexibility and might be more feasible for clinical applications. In our experiments, we found using a pseudo-3D input improves model performance compared to using a single 2D slice as input. This is reasonable as the adjacent slices contain useful spatial information for denoising the center slice. The performance improved as more adjacent slices were included as the input (N=3,5,7). Nevertheless, the improvement from 5 to 7 slices was marginal and since further away slices has less anatomically relevant information, we assume using more slices will have limited improvement to the performance. It is interesting that adding an M0 channel can improve the performance of similarity metrics and SNR to a great extent, consistent with Gong et al[8]. This may be because the M0 image provides more structural information than perfusion images. However, we also found a large bias in WM CBF with models including M0 input, which was larger than 10% of the reference values. In addition, CBF biases increased with pseudo-3D input with larger number of slices. These observations prompted us to pick pseudo-3D models with 3 slices input as the optimal choice for achieving improved SNR without affecting the accuracy in CBF quantification.

Among the 3 model backbones, a Swin Transformer-based network (SwinIR) achieved better performance than CNN-based network (ResNet and DWAN) in almost every input conditions. Though previous studies have shown Transformer-based models can perform better than CNN-only based network[15], it usually requires a large amount of training data, which can be a challenge in medical imaging. In this study, we used the SwinIR network that combined CNN and Transformer. The convolution layers can enhance the translational equivalence of the Transformer models. Also, we used a sub-volume training strategy and data augmentation techniques to increase the training data size. Given that ASL is always acquired with repetitions, we were able to obtain multiple input-reference pairs for each scan. The results may suggest that SwinIR may be ideally suited for denoising ASL images by capturing the spatial and temporal information in a 4D ASL dataset. More inference data and future studies are warranted to thoroughly evaluate Transformer-based models.

### 4.3 Model generalizability and extension to multidelay ASL

We also evaluated the model performance on 3D pCASL datasets from different cohorts acquired on 3 vendor platforms with different acquisition parameters to test the model generalizability (without finetuning). Overall, the denoising performance was moderate in new datasets, the SNR can be improved by about 50% if all repetitions were averaged by (except for GE data). The SNR can reach the level of reference with about 50% average of repetitions. Among the model backbones, SwinIR showed a moderate performance on all three test-only cohorts with the best performance in Siemens data and second place for Philips and GE data. This result supports the generalizability of Transformer and CNN-based DL models for ASL denoising.

The idea of adding input channels was extended to multi-delay ASL data. Multi-delay ASL can simultaneously measure CBF and ATT and may be more advantageous for characterizing cerebrovascular disorders compared to single-delay ASL [33]–[35]. However, since the number of repetitions were distributed across multiple PLDs, the SNR for each delay is relatively low, which makes it a desirable application for DL denoising. Traditional methods that denoise each perfusion image may disrupt the temporal relationship across PLDs, leading to bias or error in fitting perfusion parameters. In this study, we combined perfusion images in adjacent PLDs to provide a constraint for the model to preserve the dynamic signal changes for multi-delay ASL. The results showed that the CBF value was marginally affected, while ATT can be more precisely estimated with the added temporal channel (i.e., pseudo-4D input). This makes multi-delay ASL more feasible for clinical applications by reducing the total scan time to about 5 minutes.

### 4.4 Study limitation

There are several limitations for this study. First, we observed biases in fitted CBF and ATT values with DL denoising, although the differences were not large, cautions still need to be taken in clinical applications. Since existing literature reported test-retest variability of ASL scans on the order of 10%[25]–[29], we consider biases less than 10% acceptable in clinical applications. One possible solution to this may be developing direct mapping from the input to quantitative maps, like the work in[36] and this would be a direction in future studies.

Second, the training sample size of the model is still relatively small compared to other computer vision tasks. This is important for improving the generalizability of the model to make it work for ASL data of multiple vendors rather than the patterns seen in the training data. Third, we only used generally healthy subjects in the training, the performance of the proposed DL method needs to be further tested on subjects with neurologic disorders such as stroke and brain tumor. Future work may include more clinical ASL cases in training and/or combined with a fine-tuning approach.

## 5. Conclusion

In conclusion, we presented Swin Transformer-based DL model for spatial and spatiotemporal denoising of single-delay or multi-delay ASL data respectively. This may facilitate the clinical translation of ASL by reducing scan time and/or enhancing the perfusion image.

## Data Availability

All data produced in the present study are available upon reasonable request to the authors

## Acknowledgement

This work was supported by National Institute of Health (NIH) grants UF1-NS100614, R01-NS114382, R01-EB032169, R01-EB028297, UF1-NS100588, and U24-NS100591. The authors thank the MarkVCID consortium (www.markvcid.org) for collecting and sharing the multi-site ASL data.

## Data availability statement

The 3 DL models are opensource as described in their corresponding publications. The ASL datasets 1, 2 and 6 collected at the University Southern California (USC) can be requested by contacting the corresponding author and through USC Material Transfer Agreement (MTA). The multi-site ASL datasets 3, 4 and 5 can be requested by contacting the MarkVCID consortium (www.markvcid.org).

## Reference

[1] D. C. Alsop et al., “Recommended Implementation of Arterial Spin Labeled Perfusion MRI for Clinical Applications: A consensus of the ISMRM Perfusion Study Group and the European Consortium for ASL in Dementia,” 2015.

[2] S. M. Spann et al., “Robust single-shot acquisition of high resolution whole brain ASL images by combining time-dependent 2D CAPIRINHA sampling with spatio-temporal TGV reconstruction,” NeuroImage, vol. 206, p. 116337, Feb. 2020, doi:10.1016/j.neuroimage.2019.116337.

[3] J. A. Wells, D. L. Thomas, M. D. King, A. Connelly, M. F. Lythgoe, and F. Calamante, “Reduction of errors in ASL cerebral perfusion and arterial transit time maps using image de-noising,” Magn. Reson. Med., vol. 64, no. 3, pp. 715–724, Sep. 2010, doi:10.1002/mrm.22319.

[4] Y. LeCun, Y. Bengio, and G. Hinton, “Deep learning,” Nature, vol. 521, mno. 7553, Art. no. 7553, May 2015, doi:10.1038/nature14539.

[5] O. Ronneberger, P. Fischer, and T. Brox, “U-Net: Convolutional Networks for Biomedical Image Segmentation.” arXiv, May 18 2015. doi:10.48550/arXiv.1505.04597.

[6] K. He, X. Zhang, S. Ren, and J. Sun, “Deep Residual Learning for Image Recognition.” arXiv, Dec. 102015. doi:10.48550/arXiv.1512.03385.

[7] D. Xie et al., “Denoising arterial spin labeling perfusion MRI with deep machine learning,” Magn. Reson. Imaging, vol. 68, pp. 95–105, May 2020, doi:10.1016/j.mri.2020.01.005.

[8] E. Gong, J. Guo, J. Liu, A. Fan, J. Pauly, and G. Zaharchuk, “Deep learning and multi-contrast based denoising for low-SNR Arterial Spin Labeling (ASL) MRI,” in Medical Imaging 2020: Image Processing, B. A. Landman andI. Išgum, Eds., Houston, United States: SPIE, Mar. 2020, p. 21. doi:10.1117/12.2549765.

[9] K. H. Kim, S. H. Choi, and S.-H. Park, “Improving Arterial Spin Labeling by Using Deep Learning,” Radiology, vol. 287, no. 2, pp. 658–666, May 2018, doi:10.1148/radiol.2017171154.

[10] L. Zhang et al., “Improving Sensitivity of Arterial Spin Labeling Perfusion MRI in Alzheimer’s Disease Using Transfer Learning of Deep Learning-Based ASL Denoising,” J. Magn. Reson. Imaging, vol. 55, no. 6, pp. 1710–1722, 2022, doi:10.1002/jmri.27984.

[11] A. Vaswani et al., “Attention Is All You Need.” arXiv, Dec. 05 2017. Accessed: Mar. 152023. [Online]. Available: http://arxiv.org/abs/1706.03762

[12] A. Dosovitskiy et al., “An Image is Worth 16×16 Words: Transformers for Image Recognition at Scale.” arXiv, Jun. 032021. Accessed: Mar. 152023. [Online]. Available: http://arxiv.org/abs/2010.11929

[13] K. He et al., “Transformers in Medical Image Analysis: A Review.” arXiv, Aug. 19 2022. Accessed: Mar. 152023. [Online]. Available: http://arxiv.org/abs/2202.12165

[14] Z. Liu et al., “Swin Transformer: Hierarchical Vision Transformer using Shifted Windows.” arXiv, Aug. 17 2021. Accessed: Mar. 15 2023. [Online]. Available: http://arxiv.org/abs/2103.14030

[15] J. Liang, J. Cao, G. Sun, K. Zhang, L. Van Gool, and R. Timofte, “SwinIR: Image Restoration Using Swin Transformer,” in 2021 IEEE/CVF International Conference on Computer Vision Workshops (ICCVW), Montreal, BC, Canada: IEEE, Oct. 2021, pp. 1833–1844. doi:10.1109/ICCVW54120.2021.00210.

[16] S. He, H. Xue, J. Cheng, L. Wang, Y. Wang, and Y. Zhang, “Tackling the oversmoothing problem of CNN-based hyperspectral image classification,” J. Appl. Remote Sens., vol. 16, no. 4, p. 048506, Nov. 2022, doi:10.1117/1.JRS.16.048506.

[17] H. Lu et al., “MarkVCID cerebral small vessel consortium: II. Neuroimaging protocols,” Alzheimers Dement., vol. 17, no. 4, pp. 716–725, 2021, doi:10.1002/alz.12216.

[18] K. Jann et al., “Cross-Vendor Test-Retest Analysis of 3D pCASL Cerebral Blood Flow”.

[19] R. B. Buxton, L. R. Frank, E. C. Wong, B. Siewert, S. Warach, and R. R. Edelman, “A general kinetic model for quantitative perfusion imaging with arterial spin labeling,” Magn. Reson. Med., vol. 40, no. 3, pp. 383–396, Sep. 1998, doi:10.1002/mrm.1910400308.

[20] B. Lim, S. Son, H. Kim, S. Nah, and K. M. Lee, “Enhanced Deep Residual Networks for Single Image Super-Resolution.” arXiv, Jul. 10 2017. Accessed: Mar. 19 2023. [Online]. Available: http://arxiv.org/abs/1707.02921

[21] M. H. Vu, G. Grimbergen, T. Nyholm, and T. Löfstedt, “Evaluation of Multi-Slice Inputs to Convolutional Neural Networks for Medical Image Segmentation,” Med. Phys., vol. 47, no. 12, pp. 6216–6231, Dec. 2020, doi:10.1002/mp.14391.

[22] A. Paszke et al., “PyTorch: An Imperative Style, High-Performance Deep Learning Library.” arXiv, Dec. 03 2019. Accessed: Mar. 16 2023. [Online]. Available: http://arxiv.org/abs/1912.01703

[23] D. Huang, B. Wu, K. Shi, L. Ma, Y. Cai, and X. Lou, “Reliability of Three-Dimensional Pseudo-Continuous Arterial Spin Labeling MR Imaging for Measuring Visual Cortex Perfusion on Two 3T Scanners,” PLOS ONE, vol. 8, no. 11, p. e79471, Nov. 2013, doi:10.1371/journal.pone.0079471.

[24] D. A. Feinberg, A. Beckett, and L. Chen, “Arterial spin labeling with simultaneous multi-slice echo planar imaging,” Magn. Reson. Med., vol. 70, no. 6, pp. 1500–1506, Dec. 2013, doi:10.1002/mrm.24994.

[25] Y. Chen, D. J. J. Wang, and J. A. Detre, “Test-retest reliability of arterial spin labeling with common labeling strategies,” J. Magn. Reson. Imaging JMRI, vol. 33, no. 4, pp. 940–949, Apr. 2011, doi:10.1002/jmri.22345.

[26] E. Kilroy, L. Apostolova, C. Liu, L. Yan, J. Ringman, and D. J. J. Wang, “Reliability of two-dimensional and three-dimensional pseudo-continuous arterial spin labeling perfusion MRI in elderly populations: comparison with 15O-water positron emission tomography,” J. Magn. Reson. Imaging JMRI, vol. 39, no. 4, pp. 931–939, Apr. 2014, doi:10.1002/jmri.24246.

[27] D. J. Hodkinson et al., “Quantifying the test–retest reliability of cerebral blood flow measurements in a clinical model of on-going post-surgical pain: A study using pseudocontinuous arterial spin labelling,” NeuroImage Clin., vol. 3, pp. 301–310, Jan. 2013, doi:10.1016/j.nicl.2013.09.004.

[28] T. Lin, J. Qu, Z. Zuo, X. Fan, H. You, and F. Feng, “Test-retest reliability and reproducibility of long-label pseudo-continuous arterial spin labeling,” Magn. Reson. Imaging, vol. 73, pp. 111–117, Nov. 2020, doi:10.1016/j.mri.2020.07.010.

[29] K. Jann et al., “Evaluation of Cerebral Blood Flow Measured by 3D PCASL as Biomarker of Vascular Cognitive Impairment and Dementia (VCID) in a Cohort of Elderly Latinx Subjects at Risk of Small Vessel Disease,” Front. Neurosci., vol. 15, 2021, Accessed: Mar. 292023. [Online]. Available: https://www.frontiersin.org/articles/10.3389/fnins.2021.627627

[30] D. Wang, Z. Wu, and H. Yu, “TED-net: Convolution-free T2T Vision Transformer-based Encoder-decoder Dilation network for Low-dose CT Denoising.” arXiv, Jun. 08 2021. doi:10.48550/arXiv.2106.04650.

[31] A. Luthra, H. Sulakhe, T. Mittal, A. Iyer, and S. Yadav, “Eformer: Edge Enhancement based Transformer for Medical Image Denoising.” arXiv, Nov. 09 2021. doi:10.48550/arXiv.2109.08044.

[32] L. Zhang et al., “Spatial adaptive and transformer fusion network (STFNet) for low-count PET blind denoising with MRI,” Med. Phys., vol. 49, no. 1, pp. 343–356, Jan. 2022, doi:10.1002/mp.15368.

[33] X. Golay and M.-L. Ho, “Multidelay ASL of the pediatric brain,” Br. J. Radiol., vol. 95, no. 1134, p. 20220034, Jun. 2022, doi:10.1259/bjr.20220034.

[34] D. J. J. Wang et al., “Multi-delay multi-parametric arterial spin-labeled perfusion MRI in acute ischemic stroke — Comparison with dynamic susceptibility contrast enhanced perfusion imaging,” NeuroImage Clin., vol. 3, pp. 1–7, Jul. 2013, doi:10.1016/j.nicl.2013.06.017.

[35] X. Lou et al., “Multi-delay ASL can identify leptomeningeal collateral perfusion in endovascular therapy of ischemic stroke,” Oncotarget, vol. 8, no. 2, pp. 2437–2443, Dec. 2016, doi:10.18632/oncotarget.13898.

[36] N. J. Luciw, Z. Shirzadi, S. E. Black, M. Goubran, and B. J. MacIntosh, “Automated generation of cerebral blood flow and arterial transit time maps from multiple delay arterial spin-labeled MRI,” Magn. Reson. Med., vol. 88, no. 1, pp. 406–417, Jul. 2022, doi:10.1002/mrm.29193.

