## Supplementary for "Transformer based deep learning denoising of single and multi-delay 3D Arterial Spin Labeling"

Supplementary information

| Single delay | Center slice | Adjacent slices | M0 |
| --- | --- | --- | --- |
| Baseline 2D | √ | 0 | × |
| Pseudo3d 3 slices | √ | 2 | × |
| Pseudo3d 5 slices | √ | 4 | × |
| Pseudo3d 7 slices | √ | 6 | × |
| 2d with M0 | √ | 0 | √ |
| 3 slices with M0 | √ | 2 | √ |
| Multi-delay |  |  |  |
| 3 slices (spatial only) | √ | 0 | 0 |
| 3 slices + 3 PLDs  (spatiotemporal) | √ | 2 | 2 |

Supplementary Table S1. Details of the input settings in the experiment. (M0 is the proton density image)

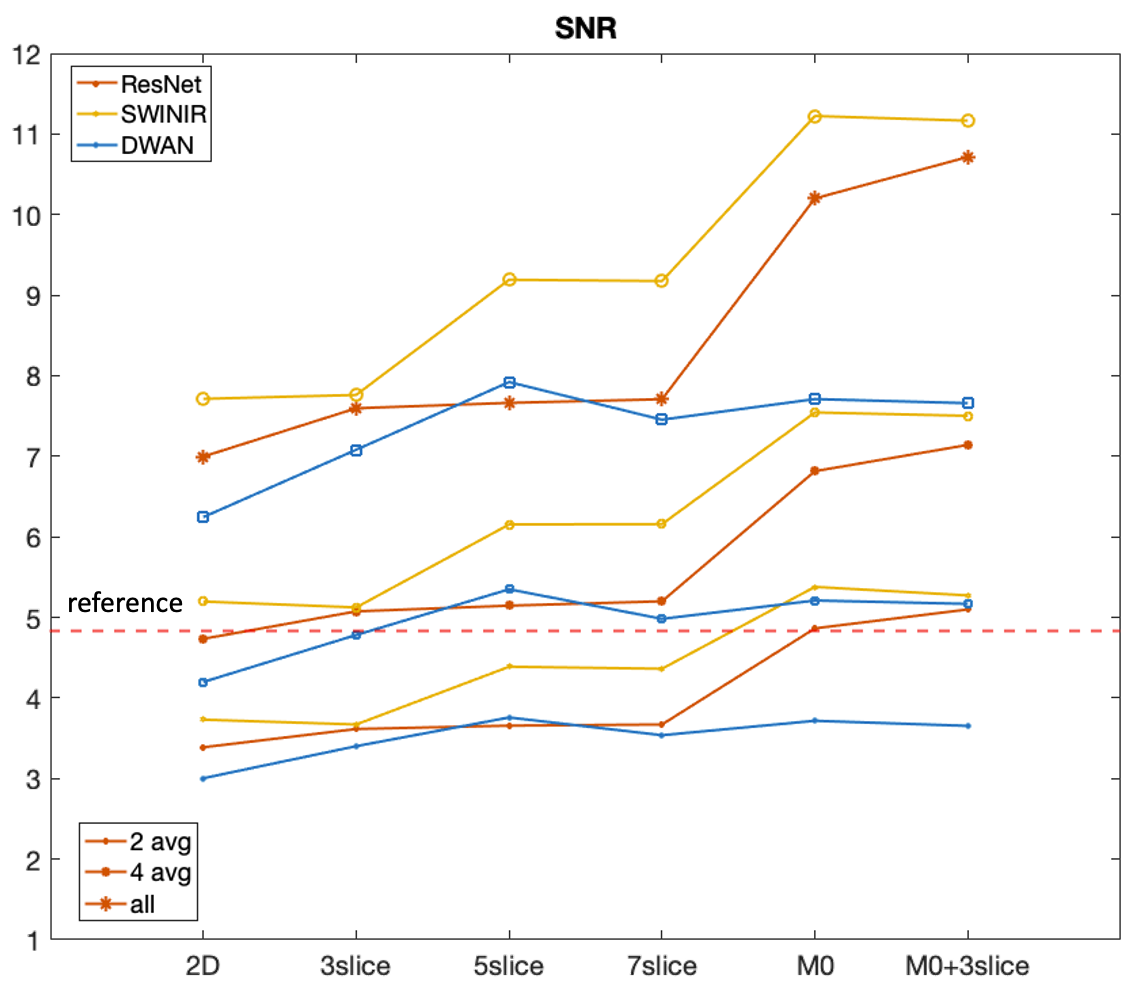

Supplement Figure S1. SNR of the output perfusion image with different proportion of averaging. For SNR, within the same proportion of averaging, the trend is similar to the similarity metrics. More averaging will improve SNR. The reference SNR (dashed red line) can be achieved by about 50% of the input and with 100% of the input, the SNR can be improved by about 2-fold.

| **Model** | **SSIM** | **PSNR** |
| --- | --- | --- |
| input | 0.876±0.019 | 22.659±1.311 |
| ResNet_2D | 0.877±0.020 | 24.045±1.359 |
| ResNet_3 | 0.883±0.020 | 24.293±1.392 |
| ResNet_5 | 0.886±0.019 | 24.495±1.421 |
| ResNet_7 | 0.885±0.019 | 24.511±1.434 |
| ResNet_2D_M0 | 0.898±0.019 | 25.718±1.480 |
| ResNet_3_M0 | 0.897±0.018 | 25.793±1.464 |
| SwinIR_2D | 0.875±0.020 | 24.320±1.385 |
| SwinIR_3 | 0.884±0.020 | 24.577±1.445 |
| SwinIR_5 | 0.893±0.019 | 25.281±1.521 |
| SwinIR_7 | 0.895±0.019 | 25.342±1.513 |
| SwinIR_2D_M0 | 0.901±0.018 | 25.903±1.483 |
| SwinIR_3_M0 | 0.902±0.019 | 25.954±1.478 |
| DWAN_2D | 0.873±0.019 | 23.591±1.425 |
| DWAN_3 | 0.881±0.019 | 24.241±1.454 |
| DWAN_5 | 0.886±0.019 | 24.704±1.495 |
| DWAN_7 | 0.882±0.017 | 24.072±1.409 |
| DWAN_2D_M0 | 0.884±0.018 | 24.705±1.448 |

Supplementary Table S2. Similarity metrics for all model backbones and input settings.

| **Model** |  | **Siemens (test data)** |  |
| --- | --- | --- | --- |
|  | **25% SNR** | **50% SNR** | **100% SNR** |
| input | 2.333±0.651 | 3.226±1.002 | 4.840±1.627 |
| ResNet_2D | 3.388±0.939 | 4.734±1.501 | 6.996±2.449 |
| ResNet_3 | 3.615±0.991 | 5.075±1.587 | 7.596±2.699 |
| ResNet_5 | 3.657±1.012 | 5.146±1.601 | 7.663±2.716 |
| ResNet_7 | 3.671±1.014 | 5.200±1.631 | 7.708±2.775 |
| ResNet_2D_M0 | 4.864±1.258 | 6.815±2.124 | 10.200±3.433 |
| ResNet_3_M0 | 5.099±1.317 | 7.141±2.266 | 10.715±3.729 |
| SwinIR_2D | 3.731±0.996 | 5.197±1.611 | 7.713±2.530 |
| SwinIR_3 | 3.671±0.944 | 5.124±1.498 | 7.671±2.502 |
| SwinIR_5 | 4.388±1.113 | 6.154±1.774 | 9.191±2.861 |
| SwinIR_7 | 4.363±1.094 | 6.158±1.765 | 9.175±2.883 |
| SwinIR_2D_M0 | 5.379±1.325 | 7.554±2.211 | 11.223±3.443 |
| SwinIR_3_M0 | 5.273±1.323 | 7.502±2.248 | 11.165±3.751 |
| DWAN_2D | 3.004±0.882 | 4.199±1.417 | 6.245±2.270 |
| DWAN_3 | 3.402±0.978 | 4.782±1.567 | 7.081±2.556 |
| DWAN_5 | 3.785±1.022 | 5.349±1.639 | 7.921±2.663 |
| DWAN_7 | 3.538±1.077 | 4.982±1.747 | 7.454±2.822 |
| DWAN_2D_M0 | 3.717±1.123 | 5.210±1.877 | 7.710±2.943 |

Supplementary Table S3. SNR with different proportion of averaging for all model backbones and input settings.

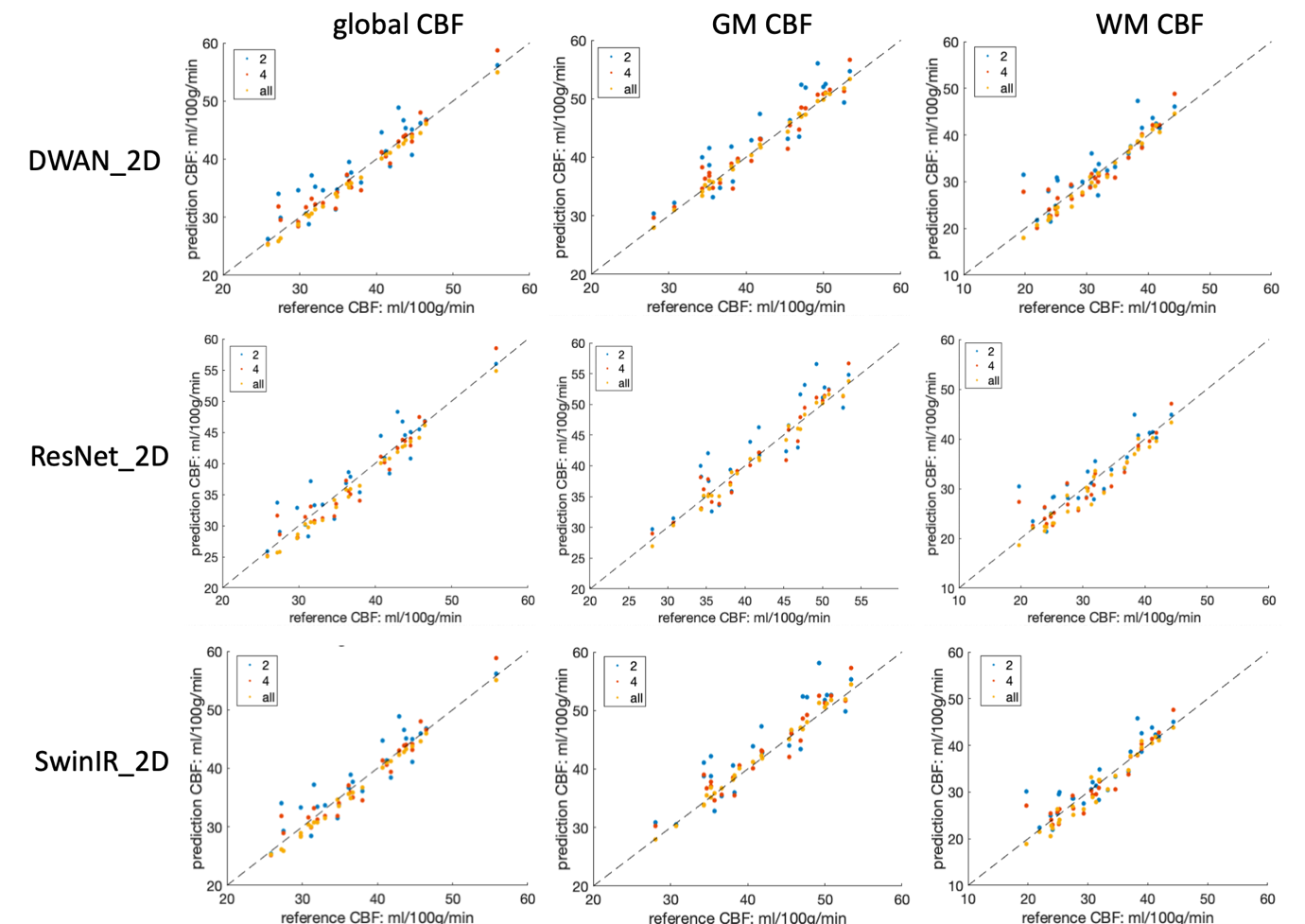

Supplementary Figure S2. Scatter plot of the predicted CBF values of whole brain, gray matter and white matter averaged with 2, 4, and all repetitions. 3 slice input condition for 3 model backbones are shown.

| **Model** | **Test data (from dataset #1-2)** | | | | | | | | | |
| --- | --- | --- | --- | --- | --- | --- | --- | --- | --- | --- |
|  | Whole brain | | | Gray matter | | | | White matter | | |
|  | Mean bias (95% CI) | In percentage | ICC | Mean bias (95% CI) | In percentage | ICC | Mean bias (95% CI) | | In percentage | ICC |
| ResNet_2D | -1.187 [-1.384,-0.989] | -3.38% [-4.06%,-2.69%] | 0.985 | -0.304 [-0.633,0.025] | -0.86% [-1.68%,-0.04%] | 0.995 | -1.371 [-1.970,-0.771] | | -4.48% [-6.45%,-2.51%] | 0.959 |
| ResNet_3 | -1.152 [-1.344,-0.959] | -3.26% [-3.90%,-2.62%] | 0.986 | 0.167 [-0.090,0.424] | 0.29% [-0.33%,0.91%] | 0.997 | -2.479 [-3.000,-1.958] | | -8.09% [-9.87%,-6.32%] | 0.923 |
| ResNet_5 | -1.011 [-1.192,-0.830] | -2.90% [-3.52%,-2.27%] | 0.989 | 0.458 [0.100,0.817] | 0.96% [0.17%,1.75%] | 0.993 | -2.306 [-2.888,-1.724] | | -7.47% [-9.39%,-5.55%] | 0.926 |
| ResNet_7 | -0.918 [-1.092,-0.743] | -2.63% [-3.23%,-2.04%] | 0.991 | 0.423 [0.031,0.814] | 0.87% [0%,1.73%] | 0.992 | -1.937 [-2.558,-1.316] | | -6.21% [-8.17%,-4.25%] | 0.937 |
| ResNet_2D_M0 | -2.078 [-2.297,-1.859] | -5.84% [-6.64%,-5.03%] | 0.959 | -0.107 [-0.659,0.444] | -0.43% [-1.74%,0.89%] | 0.987 | -5.631 [-6.267,-4.996] | | -18.15% [-20.15%,-16.15%] | 0.717 |
| ResNet_3_M0 | -1.949 [-2.191,-1.707] | -5.50% [-6.38%,-4.63%] | 0.963 | -0.071 [-0.650,0.507] | -0.40% [-1.77%,0.97%] | 0.986 | -4.943 [-5.738,-4.148] | | -15.77% [-18.14%,-13.40%] | 0.748 |
| SwinIR_2D | -0.894 [-1.062,-0.727] | -2.55% [-3.13%,-1.98%] | 0.991 | 0.336 [0.060,0.612] | 0.75% [0.09%,1.40%] | 0.996 | -0.743 [-1.366,-0.121] | | -2.76% [-4.89%,-0.63%] | 0.972 |
| SwinIR_3 | -0.877 [-1.032,-0.721] | -2.51% [-3.05%,-1.97%] | 0.992 | 0.895 [0.585,1.204] | 2.05% [1.41%,2.70%] | 0.99 | -2.417 [-2.943,-1.892] | | -8.16% [-10.17%,-6.14%] | 0.929 |
| SwinIR_5 | -0.933 [-1.100,-0.766] | -2.65% [-3.21%,-2.09%] | 0.99 | 1.175 [0.754,1.596] | 2.73% [1.79%,3.67%] | 0.983 | -2.965 [-3.597,-2.333] | | -9.59% [-11.64%,-7.53%] | 0.891 |
| SwinIR_7 | -1.025 [-1.182,-0.867] | -2.91% [-3.46%,-2.35%] | 0.989 | 1.139 [0.685,1.593] | 2.62% [1.63%,3.61%] | 0.982 | -3.425 [-4.035,-2.815] | | -11.03% [-12.98%,-9.08%] | 0.866 |
| SwinIR_2D_M0 | -1.714 [-1.943,-1.484] | -4.80% [-5.54%,-4.05%] | 0.971 | 0.875 [0.405,1.344] | 1.97% [0.83%,3.10%] | 0.985 | -6.634 [-7.381,-5.887] | | -21.44% [-23.87%,-19.01%] | 0.646 |
| SwinIR_3_M0 | -1.626 [-1.848,-1.404] | -4.54% [-5.24%,-3.83%] | 0.973 | 1.032 [0.615,1.448] | 2.37% [1.41%,3.33%] | 0.985 | -5.825 [-6.543,-5.108] | | -18.88% [-21.26%,-16.50%] | 0.705 |
| DWAN_2D | -0.840 [-0.958,-0.721] | -2.37% [-2.79%,-1.95%] | 0.993 | -0.096 [-0.290,0.099] | -0.20% [-0.65%,0.26%] | 0.998 | -0.767 [-1.185,-0.350] | | -2.98% [-4.49%,-1.47%] | 0.984 |
| DWAN_3 | -0.976 [-1.106,-0.845] | -2.74% [-3.18%,-2.30%] | 0.99 | 0.125 [-0.093,0.342] | 0.27% [-0.22%,0.76%] | 0.998 | -1.338 [-1.635,-1.041] | | -4.55% [-5.66%,-3.44%] | 0.977 |
| DWAN_5 | -0.876 [-1.020,-0.733] | -2.49% [-2.98%,-2.00%] | 0.992 | 0.611 [0.313,0.909] | 1.41% [0.75%,2.07%] | 0.993 | -1.809 [-2.257,-1.360] | | -6.01% [-7.57%,-4.46%] | 0.955 |
| DWAN_7 | -0.729 [-0.828,-0.630] | -2.05% [-2.39%,-1.70%] | 0.994 | 0.399 [0.133,0.666] | 0.97% [0.33%,1.60%] | 0.996 | -0.883 [-1.184,-0.582] | | -3.13% [-4.19%,-2.07%] | 0.987 |
| DWAN_2D_M0 | -2.056 [-2.265,-1.847] | -5.78% [-6.58%,-4.99%] | 0.96 | -1.244 [-1.803,-0.686] | -3.14% [-4.52%,-1.77%] | 0.976 | -3.707 [-4.419,-2.995] | | -12.04% [-14.30%,-9.79%] | 0.845 |
| DWAN_3_M0 | -1.569 [-1.738,-1.399] | -4.41% [-5.05%,-3.78%] | 0.976 | -0.463 [-0.880,-0.046] | -1.20% [-2.20%,-0.21%] | 0.991 | -3.051 [-3.462,-2.639] | | -10.26% [-11.97%,-8.55%] | 0.905 |

Supplementary Table S4. Bias and intraclass correlation coefficient (ICC) for whole brain, GM and WM CBF for different models in 100% averaging conditions

| **Model** | **Siemens test data (dataset #3)** | | | | | | | | | |
| --- | --- | --- | --- | --- | --- | --- | --- | --- | --- | --- |
|  | Whole brain | | | Gray matter | | | | White matter | | |
|  | Mean bias (95% CI) | In percentage | ICC | Mean bias (95% CI) | In percentage | ICC | Mean bias (95% CI) | | In percentage | ICC |
| ResNet_3 | 0.958 [-1.112,3.028] | 2.35% [-2.74%,7.45%] | 0.834 | 1.413 [0.376,2.449] | 2.00% [-2.42%, 6.42%] | 0.884 | 0.952 [-1.117,3.021] | | 2.61% [-3.19%,8.42%] | **0.793** |
| SwinIR_3 | 1.175 [0.130,2.221] | 3.22% [0.62%,5.81%] | **0.915** | 2.042 [0.527,3.556] | 3.69% [1.24%, 6.14%] | **0.931** | 2.102 [0.339,3.865] | | 6.30% [1.52%,11.08%] | 0.760 |
| DWAN_3 | 1.927 [0.351,3.504] | 5.29% [1.43%,9.14%] | 0.829 | 0.812 [-1.166,2.789] | 5.21% [1.79%, 8.63%] | 0.874 | 2.684 [0.628,4.740] | | 8.00% [2.46%,13.53%] | 0.703 |
| **Model** | **Philips test data (dataset #4)** | | | | | | | | | |
|  | Whole brain | | | Gray matter | | | | White matter | | |
|  | Mean bias (95% CI) | In percentage | ICC | Mean bias (95% CI) | In percentage | ICC | Mean bias (95% CI) | | In percentage | ICC |
| ResNet_3 | 1.346 [0.847,1.845] | 3.19% [1.60%,4.78%] | 0.985 | 2.971 [2.121,3.822] | 5.61% [3.61%, 7.60%] | 0.958 | 0.633 [-0.642,1.909] | | 2.04% [-1.75%, 5.83%] | **0.960** |
| SwinIR_3 | 1.344 [0.758,1.929] | 3.30% [1.39%,5.20%] | 0.983 | 2.304 [1.431,3.177] | 4.50% [2.50%, 6.50%] | 0.970 | 1.735 [0.526,2.945] | | 5.17% [0.93%, 9.41%] | 0.932 |
| DWAN_3 | 0.763 [0.082,1.443] | 2.04% [0.01%,4.07%] | **0.990** | 0.588 [-0.622,1.798] | 1.46% [-1.11%, 4.02%] | **0.986** | 2.612 [1.926,3.299] | | 7.18% [5.03%, 9.33%] | 0.918 |
| **Model** | **GE test data (dataset #5)** | | | | | | | | | |
|  | Whole brain | | | Gray matter | | | | White matter | | |
|  | Mean bias (95% CI) | In percentage | ICC | Mean bias (95% CI) | In percentage | ICC | Mean bias (95% CI) | | In percentage | ICC |
| ResNet_3 | 0.659 [0.317,1.001] | 1.30% [0.70%,1.90%] | 0.993 | 0.504 [-0.115, 1.124] | 0.95% [-0.18%, 2.08%] | 0.991 | -0.907 [-1.643,-0.171] | | -2.18% [-3.93%, -0.44%] | **0.973** |
| SwinIR_3 | 0.573 [0.247,0.899] | 1.12% [0.55%,1.68%] | 0.995 | 0.361 [-0.003, 0.724] | 0.66% [0.00%, 1.32%] | **0.997** | 1.123 [-0.273,2.519] | | 2.55% [-0.78%, 5.89%] | 0.942 |
| DWAN_3 | 0.316 [0.044,0.587] | 0.68% [0.12%,1.23%] | **0.998** | -1.094 [-1.536, -0.651] | -2.15% [-3.08%, -1.23%] | 0.985 | 4.016 [1.272,6.759] | | 10.01% [3.60%, 16.42%] | 0.683 |

Supplementary Table S5. CBF bias of whole brain, gray matter, and white matter of 3 models for their performance on data from different vendors. Mean difference and 95% confidence intervals are shown, both in relative values and in percentage. Intraclass correlation coefficient (ICC) is also shown in the table.

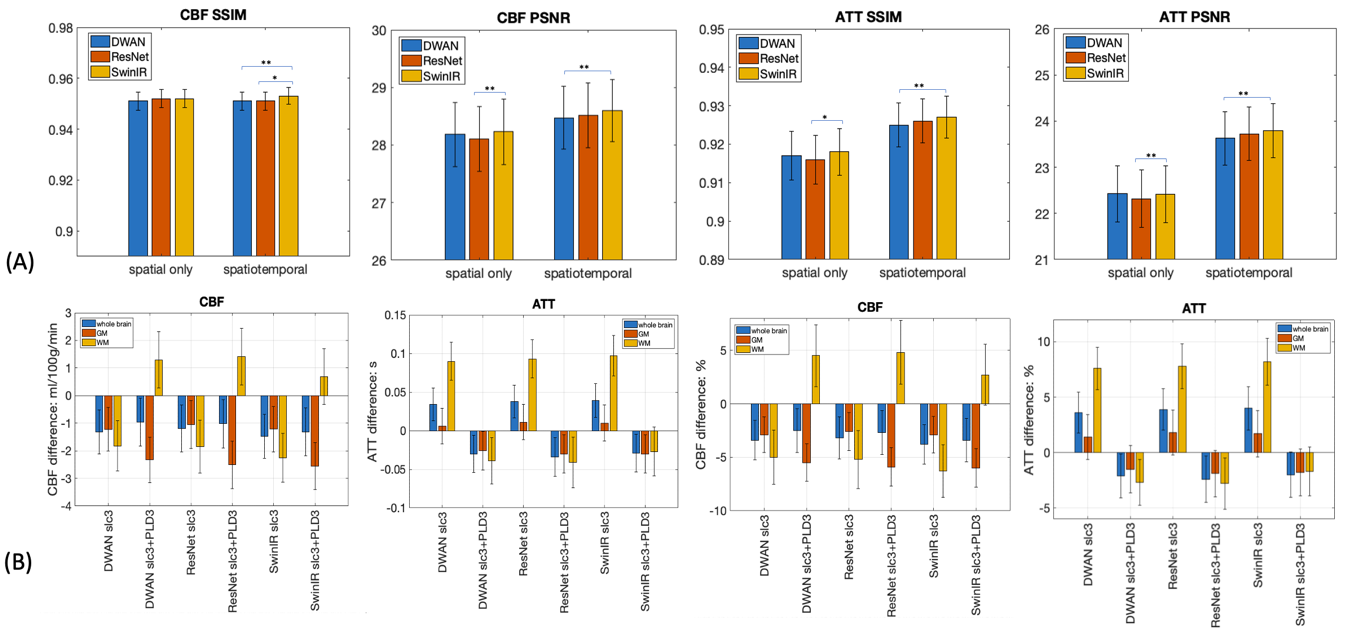

Supplementary Figure S3. Quantitative analysis of multi-delay results. (A) similarity metrics of the predicted CBF and ATT maps with reference. The performance for the CBF map is close between spatial-only and spatiotemporal denoising, but for ATT map, spatiotemporal denoising results are better than spatial-only results. (B) Bias analysis for different models. CBF difference is small for all model conditions. But for ATT, spatial-only models have much higher difference in white matter ATT. The significance of the difference was indication on the bar plot (*: p<0.05, **: p<0.01, ***: p<0.001)

| **Model** | **CBF SSIM** | **ATT SSIM** | **CBF PSNR** | **ATT PSNR** |
| --- | --- | --- | --- | --- |
| DWAN_slc3_PLD1 | 0.951±0.012 | 0.917±0.021 | 28.184±1.848 | 22.424±2.006 |
| DWAN_slc3_PLD3 | 0.951±0.012 | 0.925±0.019 | 28.475±1.793 | 23.627±1.931 |
| ResNet_slc3_PLD1 | 0.952±0.012 | 0.916±0.021 | 28.106±1.882 | 22.317±2.054 |
| ResNet_slc3_PLD3 | 0.951±0.012 | 0.92±0.019 | 28.515±1.866 | 23.723±1.919 |
| SwinIR_slc3_PLD1 | 0.952±0.012 | 0.918±0.020 | 28.230±1.888 | 22.499±2.042 |
| SwinIR_slc3_PLD3 | **0.953±0.011** | **0.927±0.018** | **28.600±1.789** | **23.791±1.933** |
| input | 0.944±0.013 | 0.905±0.025 | 27.071±1.789 | 21.749±2.151 |

Supplementary Table S6. Similarity metrics for fitted CBF and ATT maps from the multi-delay perfusion images processed by different models.

| **Model** | **CBF** | | | | | | | | | |
| --- | --- | --- | --- | --- | --- | --- | --- | --- | --- | --- |
|  | Whole brain | | | Gray matter | | | | White matter | | |
|  | Mean bias (95% CI) | In percentage | ICC | Mean bias (95% CI) | In percentage | ICC | Mean bias (95% CI) | | In percentage | ICC |
| DWAN_slc3_PLD1 | -1.318 [-3.871,1.236] | -3.39% [-9.32%,2.54%] | 0.854 | -1.224 [-3.773,1.323] | -2.88% [-8.17%,2.4%] | 0.892 | -1.825 [-4.768, 1.118] | | -5.04% [-13.24%,3.17%] | 0.744 |
| DWAN_slcl3_PLD3 | -0.959 [-3.799,1.881] | -2.52% [-9.15%,4.09%] | 0.844 | -2.331 [-5.041,0.378] | -5.46% [-11.19%,0.26%] | 0.848 | 1.298 [-2.014,4.611] | | 4.52% [-4.92%,13.97%] | 0.737 |
| ResNet_slc3_PLD1 | -1.197 [-3.909,1.516] | -3.18% [-9.48%,3.11%] | 0.852 | -1.054 [-3.839,1.731] | -2.62% [-8.36%,3.13%] | 0.886 | -1.853 [-4.916,1.209] | | -5.16% [-13.85%,3.53%] | 0.739 |
| ResNet_slc3_PLD3 | -1.022 [-3.851,1.807] | -2.70% [-9.30%,3.90%] | 0.845 | -2.511 [-5.237,0.215] | -5.88% [-11.64%,-0.11%] | 0.841 | 1.413 [-1.887,4.713] | | 4.85% [-4.69%,14.38%] | 0.746 |
| SwinIR_slcl3_PLD1 | -1.472 [-4.021,1.077] | -3.79% [-9.72%,2.15%] | 0.849 | -1.216 [-3.866,1.434] | -2.91% [-8.42%,2.59%] | 0.888 | -2.247 [-5.080,0.586] | | -6.29% [-14.14%,1.56%] | 0.737 |
| SwinIR_slcl3_PLD3 | -1.313 [-4.102,1.476] | -3.41% [-9.92%,3.11%] | 0.832 | -2.560 [-5.275,0.156] | -5.97% [-11.71%,-0.24%] | 0.835 | 0.687 [-2.523,3.898] | | 2.68% [-6.40%,11.77%] | 0.755 |
| **Model** | **ATT** | | | | | | | | | |
|  | Whole brain | | | Gray matter | | | | White matter | | |
|  | Mean bias (95% CI) | In percentage | ICC | Mean bias (95% CI) | In percentage | ICC | Mean bias (95% CI) | | In percentage | ICC |
| DWAN_slc3_PLD1 | 0.034 [-0.033,0.101] | 3.55% [-2.35%,9.45%] | 0.731 | 0.006 [-0.067,0.080] | 1.37% [-5.10%,7.85%] | 0.781 | 0.090 [0.011,0.169] | | 7.56% [1.31%, 13.8%] | 0.829 |
| DWAN_slcl3_PLD3 | -0.030 [-0.107,0.047] | -2.09% [-8.43%,4.24%] | 0.756 | -0.026 [-0.105,0.054] | -1.51% [-8.33%, 5.32%] | 0.774 | -0.039 [-0.136,0.058] | | -2.69% [-9.38%,4.01%] | 0.916 |
| ResNet_slc3_PLD1 | 0.038 [-0.030,0.105] | 3.87% [-2.11%,9.85%] | 0.692 | 0.011 [-0.062,0.084] | 1.83% [-4.66%,8.31%] | 0.769 | 0.093 [0.014,0.173] | | 7.85% [1.44%,14.25%] | 0.795 |
| ResNet_slc3_PLD3 | -0.034 [-0.114,0.046] | -2.38% [-9.01%,4.25%] | 0.716 | -0.030 [-0.109,0.049] | -1.88% [-8.61%,4.85%] | 0.772 | -0.041 [-0.146,0.064] | | -2.76% [-10.23%,4.71%] | 0.871 |
| SwinIR_slcl3_PLD1 | 0.039 [-0.031,0.109] | 4.01% [-2.19%,10.21%] | 0.690 | 0.010 [-0.065,0.085] | 1.73% [-4.88%,8.35%] | 0.765 | 0.097 [0.013,0.181] | | 8.18% [1.40%,14.97%] | 0.793 |
| SwinIR_slcl3_PLD3 | -0.029 [-0.108,0.050] | -1.96% [-8.48%,4.56%] | 0.736 | -0.030 [-0.109,0.050] | -1.85% [-8.64%,4.95%] | 0.777 | -0.027 [-0.129,0.074] | | -1.66% [-8.74%,5.42%] | 0.894 |

Supplementary Table S7. Bias and ICC for global, GM and WM CBF and ATT for multi-delay ASL quantification
